# Consensus Guideline for the Management of Peritoneal Metastases in Patients with Neuroendocrine Neoplasms

**DOI:** 10.1101/2024.04.10.24305426

**Authors:** Kiran Turaga

## Abstract

**Background:** Neuroendocrine neoplasms (NEN) with peritoneal metastases (PM) represent a complex clinical challenge due to low incidence and heterogeneous phenotypes. This manuscript describes the results of a national consensus aimed at addressing clinical management of patients with NEN-PMs.

**Methods:** An update of the 2018 Chicago consensus guidelines was conducted using a modified Delphi technique, encompassing two rounds of voting. The levels of agreement for various pathway blocks were rigorously assessed. Key systemic therapy concepts were summarized by content experts. Supporting evidence was evaluated via a rapid literature review.

**Results:** Overall, the level of evidence for the management of PM in this disease was universally low. In total, 107 participants responded in the first round, with 88/107 (82%) participating in the second round. Strong consensus (> 90%) was achieved in 5/7 (71%) and 7/7 (100%) blocks in rounds I and II respectively. A multidisciplinary approach including psychosocial and wellness assessments received a strong positive recommendation. Management of NENs with PMs was organized according to disease grade and symptom profiles. In grade 1 and 2 well-differentiated NENs, cytoreductive surgery (CRS) received strong support (>95%) following the management of functional syndromes (if present). For grade 3 well-differentiated NENs, systemic therapy is the primary recommendation, with surgical resection considered in select cases. Poorly differentiated NENs (i.e. neuroendocrine carcinomas) are predominantly treated with chemotherapy.

**Conclusion:** Given limited evidence, the consensus-driven clinical pathway offers vital clinical guidance for the management on NENs with PM. The need for high-quality evidence remains critical to the field.

## INTRODUCTION

Neuroendocrine neoplasms (NENs) are a diverse group of malignancies that arise from neuroendocrine cells throughout the body. As per the 2022 World Health Organization (WHO) classification scheme, well-differentiated NENs are defined as neuroendocrine tumors (NETs), whereas poorly differentiated NENs are distinguished as neuroendocrine carcinomas (NECs).^1^ The incidence and prevalence of NENs, including small intestine NENs, have increased nearly three-fold in the last three decades, and they are now recognized as the second most prevalent gastrointestinal neoplasm and tenth most prevalent neoplasm overall.^2,3^ The incidence of peritoneal metastasis (PM) in patients with gastroenteropancreatic neuroendocrine neoplasms (GEP-NENs) can range from 19% to 60%, most of which originate from primary midgut tumors.^4-6^

Prognosis in patients with NENs and PM (NEN-PM) varies broadly with primary tumor location, concurrent hepatic involvement, tumor grade, and patient age, with 5-year survival rates ranging from 7% to 67%.^6^ As two of the more common distant sites of disease in NENs, PM and hepatic metastases often occur concurrently.^1^ While common, the presence of PMs does not necessarily portend a poorer survival rate compared to other distant metastatic sites.^6^

In 2018, the Chicago Consensus Working Group provided multidisciplinary recommendations for the management of NEN-PMs.^7,8^ Evidence studying this disease subset remains largely retrospective and scarce. Hence, there are currently no established standard of care pathways for the management of NENs with PMs.

Herein, we present the results of a national consensus on the clinical management of patients with NENs with PM, updated in line with recent evidence synthesized through a rapid systematic review evaluating optimal management strategies for GEP-NENs with PMs.

## METHODS

This initiative was part of a national multidisciplinary consortium group process aimed at streamlining guidelines for the care of patients with peritoneal surface malignancies (PSM). The consensus and rapid review methodology has been described in detail in a separate manuscript (Submitted).^9^ Major components are summarized below.

### Consensus Group Structure

In brief, the Neuroendocrine Working Group consisting of seven neuroendocrine experts who volunteered to lead the section (PK, TH, JS, DM, AG, JK, EB). Two core group members (DS, LB) coordinated the effort. A team of six surgical residents and surgical oncology fellows conducted the rapid reviews.

### Modified Delphi Process

A modified Delphi method with two rounds of voting was employed to gather feedback regarding the clinical management pathway following preliminary synthesis of major updates since the last guideline iteration.^10^ Experts rated their agreement levels on a five-point Likert scale via a Qualtrics questionnaire. A 75% consensus threshold was set, with blocks below 90% agreement undergoing further review. Simultaneously, a summary table outlining first-line systemic therapies was generated.

### Rapid Review of the Literature

A MEDLINE search via PubMed between January 2000 and August 2023 addressed the key question: What is the optimal management strategy for abdominal NENs with peritoneal metastases? A search strategy was developed and reviewed by a medical librarian specialist, and the review protocol was pre-registered in PROSPERO (<u>CRD42023465271</u>).^11^ The Covidence platform facilitated title & abstract screening, full-text review, data extraction, and quality assessment using the Newcastle Ottawa Scale for non-randomized studies.^12-14^ The review was conducted in alignment with recommendations from the Preferred Reporting Items for Systematic reviews and Meta-Analyses (PRISMA) 2020 guidelines.^15,16^

### External Perspectives

Multiple patient advocates within the Learn Advocate Connect Neuroendocrine Tumor Society (LACNETS) reviewed the treatment pathway offered insights regarding clinical trial enrollment, research outcomes, and available resources for patients with NEN-PMs. Additionally, members of the Peritoneal Surface Oncology Group International (PSOGI) Executive Council were invited to appraise the second version of the pathway. Their comments were consolidated to evaluate alignment with global practices regarding the management of NEN-PMs.

## RESULTS

In all, 107 invited experts voted in the first Delphi round, of which 82% (n = 88) responded in the second round. Of survey responders, 72 (67%) were surgical oncologists, 18 (17%) medical oncologists, 10 (9%) pathologists, 3 (3%) radiologists, and 4 (3%) experts in other domains. Given the low quality of existing evidence in the literature, recommendations were based primarily on expert opinion. This pathway was divided into seven main blocks (Figure 1; Supplementary Figure S1) and agreement % is summarized in Tables 1-2.

**Figure 1.**
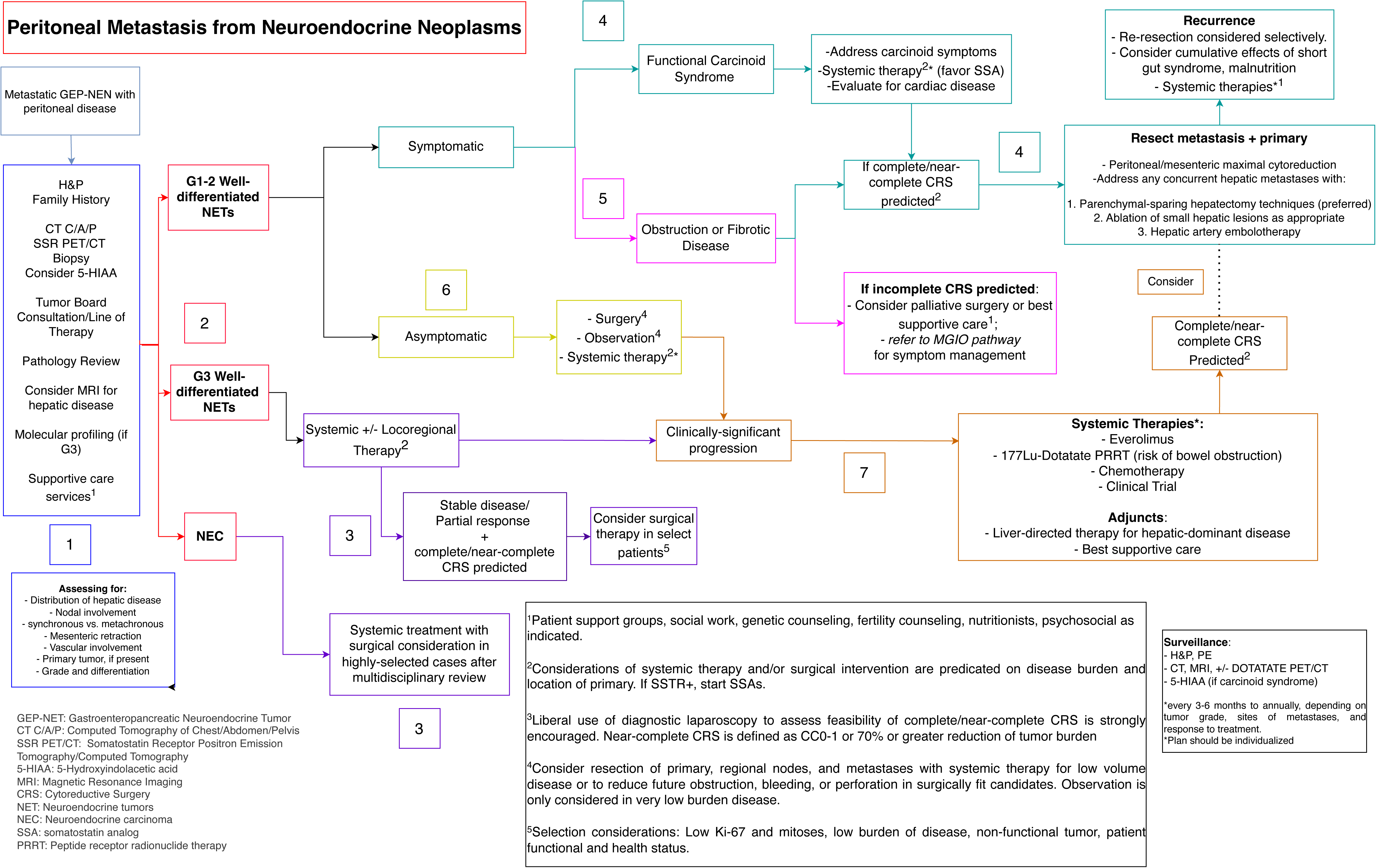

**Table 1.** Delphi Round 1 agreement table.

|  | **Strongly**<br>**agree** | **Agree** | **Neither**<br>**agree nor**<br>**disagree** | **Disagree** | **Strongly**<br>**disagree** | **Total** | **%**<br>**Agreement** |
| --- | --- | --- | --- | --- | --- | --- | --- |
| **Block 1** | 66 | 37 | 1 | 2 | 1 | 107 | 96% |
| **Block 2** | 65 | 33 | 6 | 1 | 2 | 107 | 92% |
| **Block 3** | 55 | 40 | 7 | 4 | 1 | 107 | 89% |
| **Block 4** | 64 | 35 | 5 | 0 | 3 | 107 | 93% |
| **Block 5** | 56 | 45 | 4 | 0 | 2 | 107 | 94% |
| **Block 6** | 58 | 42 | 4 | 1 | 2 | 107 | 93% |
| **Block 7** | 51 | 41 | 8 | 3 | 4 | 107 | 86% |

**Table 2.** Delphi Round 2 agreement table.

|  | **Strongly**<br>**agree** | **Agree** | **Neither**<br>**agree nor**<br>**disagree** | **Disagree** | **Strongly**<br>**disagree** | **Total** | **%**<br>**Agreement** |
| --- | --- | --- | --- | --- | --- | --- | --- |
| **Block 1** | 76 | 11 | 1 | 0 | 0 | 88 | 99% |
| **Block 2** | 78 | 8 | 1 | 0 | 1 | 88 | 98% |
| **Block 3** | 73 | 11 | 3 | 0 | 1 | 88 | 95% |
| **Block 4** | 73 | 13 | 2 | 0 | 0 | 88 | 98% |
| **Block 5** | 75 | 11 | 1 | 1 | 0 | 88 | 98% |
| **Block 6** | 75 | 10 | 2 | 0 | 1 | 88 | 97% |
| **Block 7** | 74 | 13 | 1 | 0 | 0 | 88 | 99% |

A rapid review evaluating optimal management strategies in GEP-NENs with PMs revealed 2023 abstracts as detailed in the PRISMA flow diagram (Figure 2). Twenty-three full texts were reviewed, and 9 studies were included for data extraction and quality assessment and cited in relevant sections of the document (Table 3; Supplementary Table S1-S2).^4,5,17-23^

**Figure 2:**
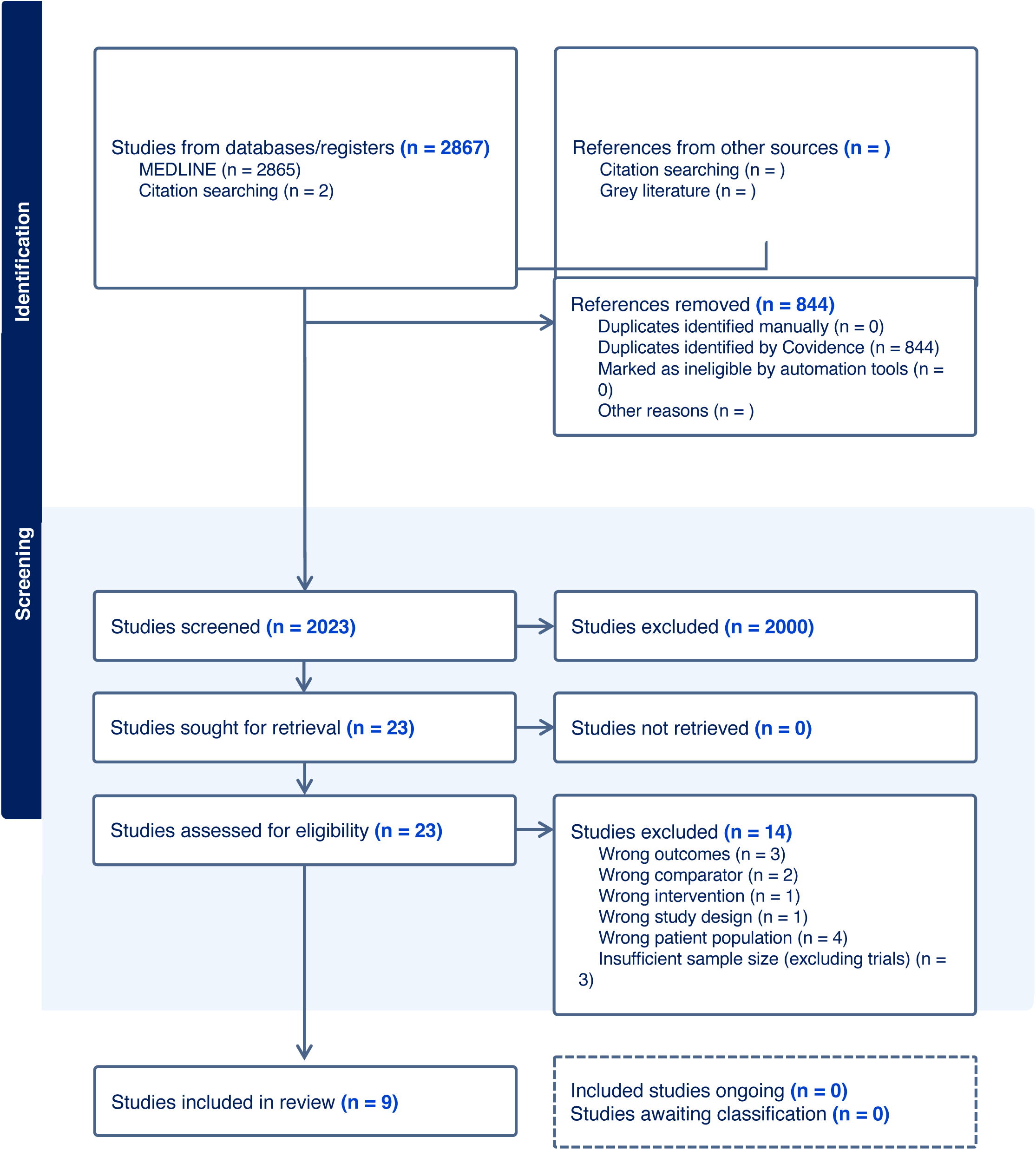
PRISMA Flow Diagram for Study Selection for Rapid Review.

**Table 3.** Description of Studies from Rapid Review.

| Author | Year Published | Country | Study Aim | Study design | Start Date | End Date | Study Population | Number of patients | Outcomes |
| --- | --- | --- | --- | --- | --- | --- | --- | --- | --- |
| Benhaim et al. [17] | 2020 | France | To evaluate if surgical resection could lead to prolonged survival for patients with peritoneal metastases | Single-institution, retrospective | 1995 | 2017 | Patients with well-differentiated NETs | 88 (PM=12, LM=41, LM+PM=35) | No difference in 5-year OS rate between groups [81% (60–100) PM vs. 78% (65.2– 92.8) LM vs. 72% (58.7–89.7) PM + LM ($p = 0.71$)]; No difference in 3-year DFS rate between groups [12.5% (2.3–68.2) PM, vs. 26.9% (16.1–45.1) LM vs. 32.4% (19.9–52.6) PM + LM ($p = 0.45$)]; Patients with SB origin had improved OS/DFS than pancreas |
| Boer et al. [18] | 2018 | Netherlands | To evaluate outcomes of palliative surgery for acute MBO | Single-institution, retrospective | 2005 | 2017 | Patients with acute obstruction caused by peritoneal carcinomatosis who underwent palliative surgery | 30 with NETs (148 overall) | NETs - improved OS (median 562 days, iQR 133-1788) and greater remaining life spent outside of hospital after discharge compared to other tumor types. |
| Elias et al. [19] | 2014 | France | To report the results of complete CRS of peritoneal metastases from NET and compare patients treated with or without HIPEC | Single-institution, retrospective | 1994 | 2012 | Patients who underwent macroscopically complete resection of well-differentiated NETs with slow tumor growth under systemic treatment (at least a 20% increase in tumor burden in the preceding 12 months) | 41 (28 CRS-HIPEC, 13 CRS alone) | 5- & 10-year OS 69% & 52%, respectively - no difference between CRS-HIPEC and CRS alone groups; 5- & 10-year DFS 17% and 6%, respectively - improved in HIPEC group in univariable analysis (fewer lung and bone metastases); no difference in major morbidity between groups |
| Elias et al. [20] | 2005 | France | To assess the prognostic impact of palliative surgery in NET-PMs | Single-institution, retrospective | 1993 | 2003 | Well-differentiated endocrine carcinoma patients (WHO 2000 Classification); two groups according to extent of resection: 1) complete resection of macroscopic peritoneal disease not possible (patients only underwent palliative debulking), 2) maximal CRS with immediate HIPEC | 37 (20 palliative surgery, 17 complete CRS) | 5-year OS 66.2% complete CRS-HIPEC vs 40.9% palliative surgery; grade 3-4 morbidity higher in CRS-HIPEC group; CRS-HIPEC group - 41% patients were disease-free right after surgery and 10% disease-free 5 years postop |
| Hajjar et al. [21] | 2022 | International Cohort | To evaluate the addition of HIPEC to CRS | Multicenter retrospective | 2002 | 2016 | Patients with small bowel-NET with PMs who underwent CRS or CRS-HIPEC | 67 (36 CRS-HIPEC, 31 CRS alone) | No difference in 5-year OS between CRS-HIPEC and CRS-alone (74.5% vs. 91.6%, $p = 0.32$). No difference in DFS in CRS-HIPEC and CRS-alone groups (30.8% vs. 0%, $p = 0.78$). CRS-HIPEC group had higher G3/G4 adverse events compared to CRS alone (>50% vs 3.4%, $p < 0.001$). |
| Merola et al. [5] | 2019 | Germany | To describe the clinical impact of PM and to report the effectiveness of available treatments in PM control | Single-institution, retrospective | 1993 | 2016 | Patients with sporadic GEP-NENs with PMs and a minimum of 12-month follow up after PM detection. | 135 (43 peritonectomy, 90 SSAs, 32 PRRT, 50 chemotherapy, 18 everolimus, 1 sunitinib) | First line therapy of SSAs demonstrated improved PFS survival (median: 114 months) compared to PRRT (25 months), everolimus (35 months), chemotherapy (13 months). Peritonectomy was associated with best PFS (152 months) compared to any other first line intervention ($p < 0.01$). Overall no difference in RFS between complete, partial, or in combination with HIPEC peritonectomies ($p = 0.45$). 22.2% of patients developed bowel obstruction at a median of 16 months after the diagnosis of PM. |
| Wonn et al. [4] | 2020 | United States | To determine how much cytoreduction needed for small bowel NETs with PMs and LMs can be achieved and corresponding survival benefits of different levels of clearance | Single-institution, retrospective | 2008 | 2019 | Patients with histologically confirmed NETs | 98 (complete CRS 49, incomplete CRS 49) | Improved OS with complete vs. incomplete CRS (OS 135 vs 51 mo, $p = 0.03$); Within incomplete CRS group, pts with localized nodules < 5 mm had improved OS vs. > 5mm size nodules (76 vs 32 mo, $p = 0.04$); patients who underwent cytoreduction for MPD and LM had longer survival (76.5 mo) compared to other groups: patients with extensive residual PM+LM cytoreduced at least 70% (39 mo), MPD+LM not cytoreduced (22 mo), extensive peritoneal disease and LM not cytoreduced (20 mo) |
| **Wright et al. [22]** | 2019 | United States | To clarify prognostic significance of peritoneal metastasis in midgut NET patients | Single-institution, retrospective | 1991 | 2017 | Patients with midgut NET with PMs who underwent surgical resection | 219 overall (80 LM; 14 PM, 53 PM + LM, 70 no mets) | Median OS 156 mo LM only vs. 133 mo PM only vs. 82 mo PM + LM (p < 0.01). Patients with PM had T4 primary lesions more frequently than those with LM only or no distant metastases. |
| **Strosberg et al. [23]** | 2021 | United States | To investigate the incidence of PRRT-induced bowel obstruction | Single-institution, retrospective | 2018 | 2019 | Patients with NET & mesenteric or peritoneal disease who received 177Lu-DOTATATE | 159 overall (81 PM/mesenteric disease) | 3% of all patients, 22% of high risk patients (extensive peritoneal or desmoplastic changes associated with mesenteric tumors) developed bowel obstruction within three months of PRRT. All patients with documented obstruction demonstrated symptomatic improvement after initiation of corticosteroid treatment. |

CRS: cytoreductive surgery; NET: neuroendocrine tumor; PM: peritoneal metastasis; LM: liver metastasis; PRRT: peptide receptor radionuclide therapy; SSA: somatostatin analogue; HIPEC: hyperthermic intraperitoneal chemotherapy; GEP-NEN: gastroenteropancreatic-neuroendocrine neoplasms; MPD: minimal peritoneal disease; OS: overall survival; RFS: recurrence-free survival; PFS: progression-free survival; DSS: disease-specific survival

**Table 4.**
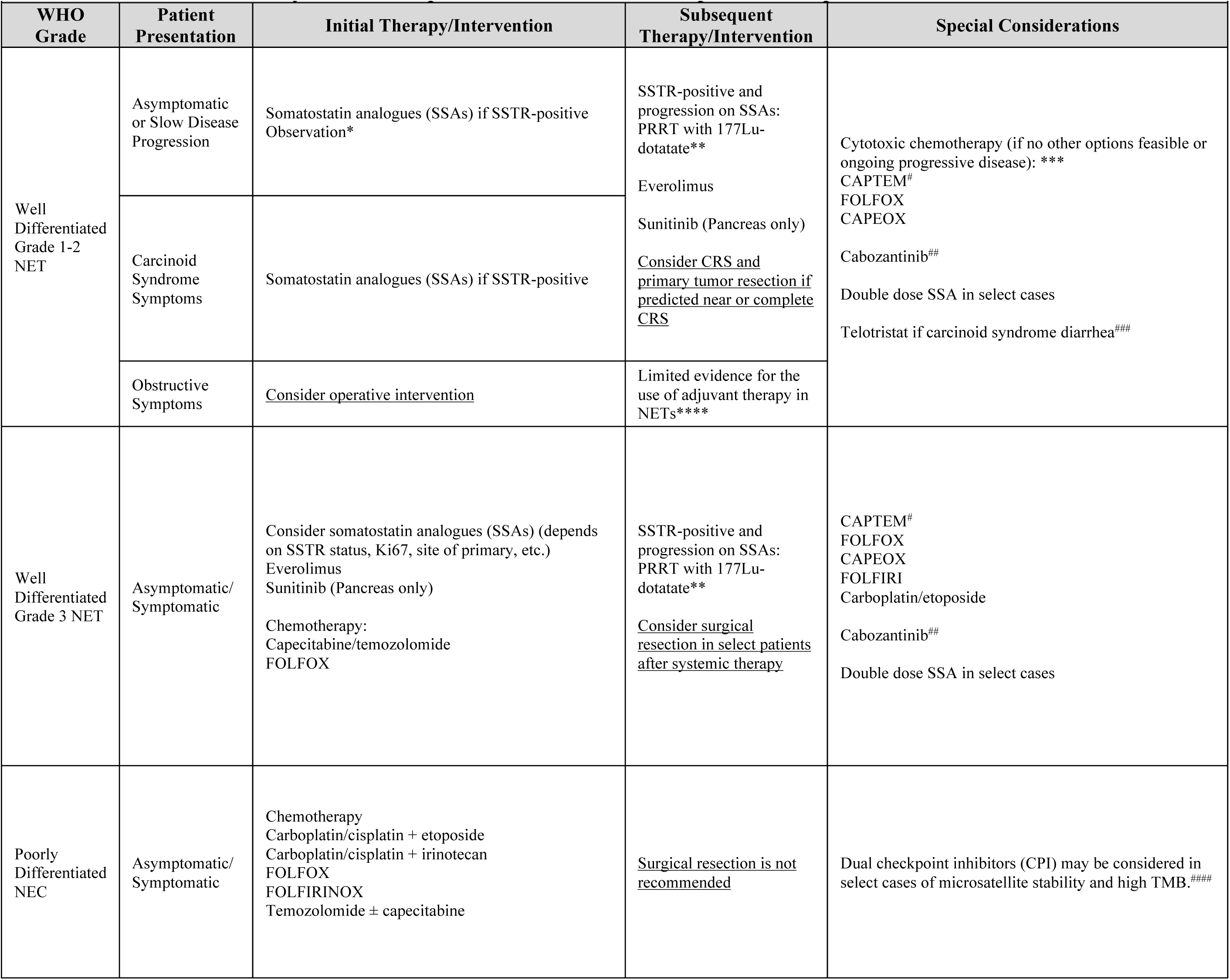
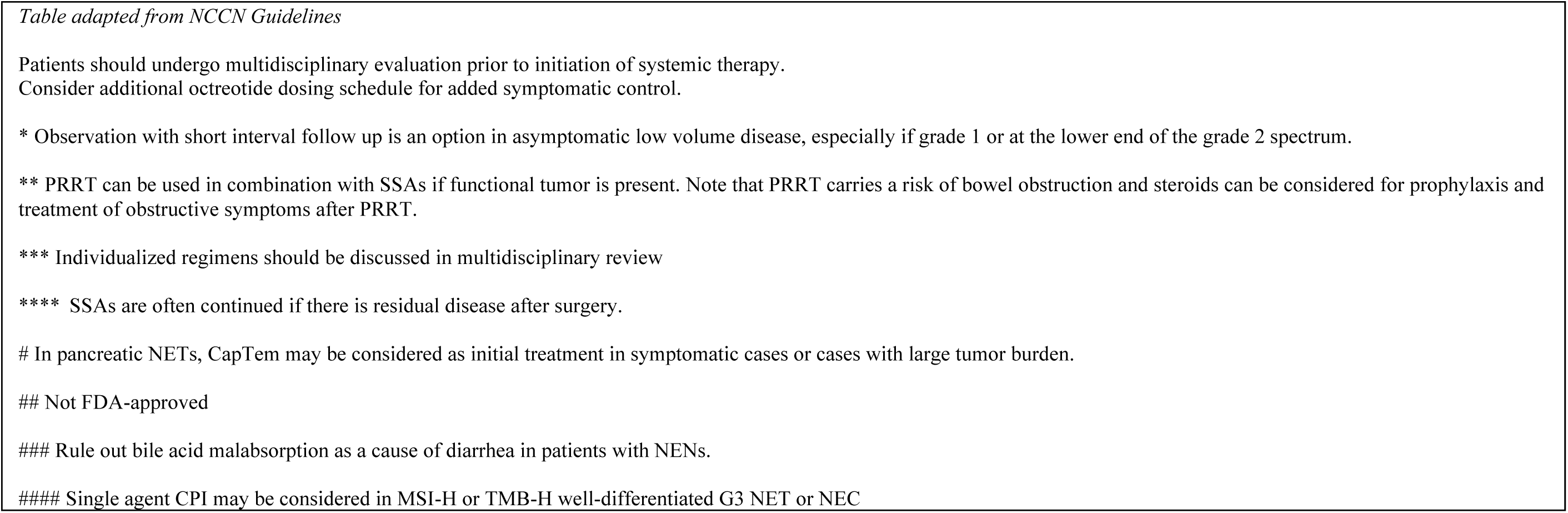
Systemic therapies in neuroendocrine neoplasms with peritoneal metastases.

### Summary of Major Updates

Building upon the 2018 Chicago Consensus Guideline, the current approach involves a more stringent consensus and review methodology while engaging a larger spectrum of experts and patient advocates.^7,8^ The updated guidelines highlight a comprehensive preoperative evaluation including psychosocial support, nutrition, fertility considerations, and linkage with patient advocacy groups. Steering away from the prior classification based primarily on liver involvement, the updated pathway draws attention to NEN-PMs based on disease grade, symptomatology, and tumor burden. The management of grade 1 and 2 well-differentiated NETs is segregated based on symptom profiles, addressing carcinoid syndrome, as well as obstruction in the setting of carcinoid-associated mesenteric fibrosis or carcinomatosis. For well-differentiated grade 3 GEP-NETs, a relatively new World Health Organization (WHO) class with limited treatment data, the primary approach leans towards systemic therapy.^1^ Surgery can be considered in patients with disease amenable to near-complete cytoreduction. Meanwhile, medical management with cytotoxic chemotherapy is recommended for NECs due to their aggressive nature and low likelihood of clinical benefit from surgical intervention.

### Block-Based Recommendations

#### Block 1: Preoperative considerations

**Agreement**: Round 1 - 97%; Round 2 - 97%

Preoperative evaluation includes a thorough examination of history and physical findings, diagnostic workup, and a multidisciplinary tumor board discussion.^24-26^ This includes a detailed assessment of functional symptoms, such as carcinoid syndrome and carcinoid heart disease. Additionally, we emphasize the early involvement for patient support groups, social work, genetic counseling, fertility specialists, psychosocial support, and programs dedicated to exercise and nutritional wellness for preoperative optimization.^27,28^

Molecular profiling of NENs, while not routinely performed for diagnostic purposes, may be helpful in distinguishing cases of G3 NETs from NECs.^1,29-31^ The role of germline testing is evolving but remains controversial, as it has traditionally only been offered to patients with a high clinical suspicion for inherited endocrine syndromes, such as multiple endocrine neoplasia, tuberous sclerosis, neurofibromatosis, or von Hippel Lindau syndrome. These germline alterations make up a very small percentage of GEP-NENs and occur more frequently in pancreatic NENs.^32-35^

The recommended imaging modalities are multiphasic CT or MRI of the abdomen and pelvis and somatostatin-receptor (SSTR)-based PET imaging, which may be either SSTR-PET/CT or SSTR-PET/MRI using ^68^Ga-DOTATATE or ^64^Cu-DOTATATE.^36^ In the presence of liver metastases, MRI imaging is considered the modality of choice due to high signal intensities observed on T2-weighted imaging.^37^ Overall, PMs remain difficult to characterize radiographically due to small tumor volume, multifocality, and tissue density, but may be more apparent on DOTATATE PET scans.^38^ Assessment of metastatic burden remains critical in the preoperative evaluation as this influences overall surgical candidacy.

#### Block 2: Stratification by grade

**Agreement**: Round 1 - 92%; Round 2 - 96%

Grading of GEP-NENs is classified by proliferation rate using the Ki-67 index and the mitotic count. Well-differentiated disease is stratified into three grades (Grade 1, 2, and 3): grade 1 NETs have a mitotic rate < 2 mitoses/2 mm^2^ and a Ki-67 index < 3%, grade 2 NETs have a mitotic rate 2-20 and/or Ki-67 index of 3-20%, and grade 3 NETs have a mitotic rate >20 or Ki-67 index >20%. NECs are molecularly and histologically distinct from well-differentiated NETs and are highly aggressive, demonstrating a Ki-67 index > 20%; however, usually above 50%.^1^

In our consensus-based pathway, peritoneal involvement in neuroendocrine neoplasms (NENs) is stratified based on histopathological grading: Grade 1/2 NET, Grade 3 NET, and NEC. This categorization aligns with the WHO system and is pivotal in clinical decision-making and prognostic estimations.^1^

#### Block 3: Grade 3 GEP-NETs and NECs

**Agreement**: Round 1- 87%; Round 2 - 96%

Well-differentiated grade 3 GEP-NETs (NET-G3) is a relatively new category in the 2017 and 2019 WHO classifications.^1,39^ The sequence of surgical and systemic therapy offered in NET-G3 with PMs is more nuanced as many prior surgical trials and studies had mixed NEC and NET-G3.^40,41^ Furthermore, many surgical studies focus on non-metastatic NET-G3 or do not subgroup by PM metastases.^42,43^ While locoregional or resectable NET-G3 disease may be treated with resection along with regional lymphadenectomy, NET-G3 tumors with PMs are most commonly offered systemic therapy potentially combined with liver-directed locoregional therapies as the initial step.^42,44^ If the disease does not progress on systemic therapy and near complete cytoreductive surgery is predicted, CRS may be offered in select patients.^45^ In cases with progression, further systemic and adjunctive therapies should be prioritized, and CRS may be considered in select cases in conjunction with multidisciplinary consultations.

NECs are an aggressive subset of NENs and are primarily treated with cytotoxic chemotherapy such as platinum/etoposide.^46,47^ Surgical intervention may be offered in limited instances based on disease stage, primary tumor site, the patient’s overall health status, clinical symptoms, and response to systemic therapy after multidisciplinary discussion.

#### Block 4: Carcinoid syndrome in lower-grade (G1/G2) GEP-NETs with PMs

**Agreement**: Round 1 - 90%; Round 2 - 96%

Patients with surgically treatable low-grade GEP-NETs should be evaluated for the carcinoid syndrome, which may occur in up to 30% of patients with liver metastases, predominantly from small bowel primary tumors.^48,49^ Carcinoid heart disease can occur in up to half of patients with carcinoid syndrome and is often characterized by plaque-like fibrous buildup on endocardial surfaces or valvular leaflets.^50^ Evaluation for carcinoid heart disease should be performed in all patients with carcinoid syndrome or evidence of serotonin production as manifested by elevated 5-HIAA in blood or urine. Other functional neuroendocrine abdominal malignancies (e.g., Zollinger-Ellison syndrome, insulinoma syndrome, etc) tend to occur in association with pancreatic primary tumors and tend not to be associated with fibrotic complications such that their PM management is similar to that of patients with non-functional NENs.

In grade 1 and 2 NETs, distinctions are made for symptomatic patients, focusing on the differentiation between carcinoid syndrome and other (functional or non-functional) tumors. When present, symptoms from carcinoid syndrome should be addressed initially, and patients started on an SSA.^51^ All at-risk patients should be monitored for cardiac disease. If either carcinoid syndrome is present or there is evidence of serotonin overproduction, patients should undergo a transthoracic echocardiogram for further evaluation.^52^ Short-acting octreotide to prevent carcinoid syndrome prior to cytoreductive surgery may be considered but its efficacy remains controversial.^53-55^

Symptomatic advanced carcinoid heart disease is most effectively treated with valvular surgery.^50^ In the setting of carcinoid heart disease, CRS, including hepatic resection, is delayed until after valve replacement occurs due to the potential congestive hepatopathy in the setting of right-sided heart failure.^51^

#### Block 4: Cytoreductive surgery in G1/G2 NET-PMs

In the treatment of low-grade well-differentiated NETs, we recommend that cytoreductive surgery be considered, targeting a 70% threshold in the reduction of tumor burden.^4,56,57^ This can be coupled with, or independent of, somatostatin analogues (SSAs). A significant observation is that most tumor bulk is usually hepatic rather than peritoneal. Hence, an open question remains on how the distribution of disease (hepatic vs. peritoneal metastases) might influence resection, current evidence for which remains equivocal. In 2020, *Benhaim et al.* evaluated 88 patients with metastatic NET, who underwent resection of PM, liver metastases, or both, and noted comparable disease-free survival (DFS) in the PM only group when compared to the liver only or the combined group.^17^ Those with small bowel primaries demonstrated longer DFS and overall survival (OS) than other primary origins.^17^ Another study retrospectively compared hepatic and peritoneal metastases in patients with small intestinal NETs and revealed that PM, either alone or in combination with hepatic metastasis, was associated with a worse prognosis compared to hepatic metastasis alone.^22^ Therefore, this preliminary work suggests PM resection may have a positive prognostic effect; however, high-level evidence on debulking remains limited, and further studies are needed.

When evaluating surgical interventions, simultaneous resection of the primary tumor and metastases is preferred if near complete CRS is feasible and primary disease is resectable. This strategy often involves addressing hepatic metastases through adjunct methods like parenchymal-sparing hepatectomy or ablation. A retrospective study showed that in patients with small bowel NETs, complete CRS was associated with improved survival compared to partial or no CRS.^4^ Moreover, patients with minimal residual disease showed better survival outcomes than those with more extensive residual disease.^4^ Particularly notable was the finding that patients with minimal peritoneal disease and significant cytoreduction of liver metastases (> 70%) had the longest survival, signifying the importance of cytoreduction in improving survival outcomes for eligible patients.^4^ In patients with unresectable distant metastases, the role of resection of asymptomatic primary tumor is controversial, but few studies have shown improved overall and progression free survival with this strategy.^58-60^

Currently, hyperthermic intraperitoneal chemotherapy (HIPEC) is generally not used in the treatment of NEN-PMs as it has limited evidence available and has been associated with a higher toxicity profile.^19,21^ A single-institution series by Elias et al compared the use of CRS with and without HIPEC in 41 patients with NEN-PMs.^19^ Overall survival of both treatment groups were similar, although the HIPEC group had an improved DFS rate. However, the authors attributed this difference to increased extra-abdominal metastases in the non-HIPEC group.^19^ Another similar study on 67 patients diagnosed with small bowel NETs who underwent CRS with or without HIPEC showed that the CRS-HIPEC group experienced a higher rate of grade III-IV adverse events (>50%) compared to only 3.4% in the CRS alone group.^21^ Furthermore, there were no added benefits in 5-year OS between the CRS-HIPEC and CRS alone groups (91.6% vs 74.5%), nor was there a difference in DFS between the two groups.^21^

In accordance with National Comprehensive Cancer Network (NCCN), North American Neuroendocrine Tumor Society (NANETS), and European Neuroendocrine Tumor Society (ENETS) guidelines, resection of the primary tumor for metastatic NETs should strongly be considered at the time of CRS to avoid locoregional complications, halt ongoing metastatic shedding, and potentially improve survival.^32,61,62^

#### Block 5: Bowel obstruction in the setting of carcinomatosis or mesenteric fibrosis

**Agreement**: Round 1 - 93%; Round 2 - 97%

Bowel obstruction is a frequent complication in patents with NEN-PM, occurring in 3-28% of cases, as highlighted in studies by Merola et al and Strosberg et al. ^5,23,63^ It is often associated with mesenteric fibrosis and necessitates surgical intervention when feasible. In instances where near-complete cytoreduction is not possible, palliative surgery or comprehensive supportive care may be considered based on individual performance status, disease biology, metastatic disease burden, and symptom profile (i.e. abdominal pain, recurrent bowel obstruction, malnutrition). These nuances are detailed in our guideline for malignant gastrointestinal obstruction in patients with peritoneal carcinomatosis.^64^ Palliative surgical correction for acute obstruction may yield lasting improvements in survival and quality of life, as demonstrated by De Boer and colleagues. In their institutional review of 135 patients with acute obstruction, 30 patients with NENs experienced longer overall survival (median 562 days, IQR 133-1788) and spent a higher proportion of their remaining life in the hospital (6.9%, IQR 1.3-21.9) compared to patients with other primary tumors.^18^ Additionally, palliative resection of the primary tumor is recommended in the setting of small intestinal NETs with a high likelihood of anticipated bowel obstructions and bleeding, potentially associated with a survival benefit.^65^

#### Block 6: Asymptomatic low-grade NETs

**Agreement**: Round 1 - 94%; Round 2 - 95%

In asymptomatic patients with limited disease, surgical resection of the primary tumor, regional lymph nodes, and distant metastases is usually recommended after multidisciplinary discussion. This approach may be combined with systemic therapies, either to manage low volume disease or to prevent complications such as obstruction, bleeding, or perforation in candidates suitable for surgery.^66,67^ The choice between systemic therapy and surgical intervention depends largely on the extent of disease burden and primary site. Since SSTR positivity is ubiquitous, initiating treatment with somatostatin analogues is often considered at diagnosis (particularly if progression has been documented), but in some cases, a purely observational approach can be adopted for those presenting with low-moderate volume asymptomatic disease.

#### Block 7: Clinical progression of disease

**Agreement**: Round 1 - 84%; Round 2 - 97%

In cases of clinically significant progression or recurrence, asymptomatic patients with grade 1/2 NETs or those with well-differentiated grade 3 NETs are usually treated with systemic therapy.^32^ This may be combined with liver-directed treatment for hepatic-dominant disease. The choice of treatment modality varies depending on the characteristics of the primary tumor and is detailed in the section below, “Principles of Systemic Therapy”. For patients who demonstrate further progression despite systemic therapy, CRS may be considered in very select patients considered amenable to complete or near complete cytoreduction, characterized as a CC0-1 resection of peritoneal metastases and successful removal of over 70% of hepatic metastases.

Surveillance for NENs typically consist of a routine history and physical examination followed by appropriate imaging studies (cross-sectional CT or MRI abdominopelvic scans with PET-CTs as clinically indicated). The recommended surveillance frequency varies by institution but typically involves follow-up every three months for NENs with unfavorable/concerning disease and every six to twelve months for resectable, locoregional disease with favorable characteristics.^68-70^

### Principles of Systemic Therapy

The following recommendations pertain primarily to well-differentiated midgut NETs (distal small intestine and proximal colon), which account for most PMs.^5^

### Somatostatin Analogs

Somatostatin analogs (SSAs) are typically the first-line systemic treatment for metastatic well-differentiated NETs due to their proven inhibitory effect on tumor growth and favorable safety profile.^71^ SSAs are also effective at palliating carcinoid syndrome. Nearly all midgut NETs express somatostatin receptors which are the target for SSAs.^72^ The phase III PROMID trial compared octreotide long-acting release (LAR) to placebo in 85 patients with well-differentiated metastatic midgut NETs. Median time to progression (TTP) was 14.3 months in the group randomized to receive octreotide-LAR versus 6 months in the placebo group.^73^ Similarly, the CLARINET study reported a significant improvement in progression-free survival (PFS) in patients with advanced, well-differentiated grade 1 and grade 2 GEP NETs (Ki-67<10%) treated with lanreotide (median PFS not reached at time of data analysis) compared with placebo (median PFS 18 months).^74^ Peritoneal metastases were not directly addressed in either study. SSAs have not been compared to placebo in tumors with KI-67 >10%. Objective response rates with SSAs are generally negligible.

### Everolimus

The RADIANT 2 study investigated everolimus plus octreotide versus placebo plus octreotide in patients with metastatic NETs and history of carcinoid syndrome (primarily midgut primaries).^75^ While there was a trend towards improvement in progression free survival (PFS), this did not meet statistical significance. The RADIANT 4 study evaluated everolimus versus placebo in patients with GI and lung NETs lacking carcinoid syndrome, a more aggressive population than studied in RADIANT 2.^76^ This study met its endpoint of significant PFS improvement (11.0 months vs. 3.9 months, p < 0.001). Objective response rate in this study was only 2%.^76^

Despite the lack of statistically significant PFS benefit of everolimus in the RADIANT 2 study, guidelines generally endorse use of everolimus with progressive GI and lung NET, even among patients with history of carcinoid syndrome.^77^ However, the benefits and risks need to be carefully evaluated: adverse effects include oral stomatitis, pneumonitis, diarrhea, immunosuppression and hyperglycemia. There is no data focusing on the role of everolimus among patients with PM.

### Sunitinib

Sunitinib was evaluated in 171 patients with advanced, well-differentiated pancreatic NETs, and it was found that the median PFS was 11.4 months in the sunitinib group and 5.5 months in the placebo group.^78^ In this study, 95% of patients had liver metastases and the objective response rate was 9.3% in the sunitinib group, but no data were provided on study drug efficacy according to liver or peritoneal tumor burden. Given the lack of evidence, sunitinib is not considered as a treatment option for patients with PM. Additionally, the use of VEGF inhibitors, such as sunitinib, carry an increased risk of perforation and bleeding.^79^

### Peptide Receptor Radionucleotide Therapy (PRRT) with 177Lu-DOTATATE

The NETTER-1 trial studied the benefit of ^177^Lu-Dotatate in patients with progressive, well-differentiated, metastatic midgut NETs. The study included 229 patients, 116 in the ^177^Lu-Dotatate group (^177^Lu-Dotatate plus 30 mg intramuscular octreotide LAR) and 113 in the control group (60 mg intramuscular octreotide LAR). The median PFS was not reached in the ^177^Lu-Dotatate group and was 8.4 months in the octreotide LAR group at the time of primary data analysis. The objective response rate was 18% in the ^177^Lu-Dotatate group and 3% in the control group. It should be noted that most patients had metastases to the liver or lymph nodes. However, 7% of the patients had PM (7 in the treatment group and 10 in the control group).^80^ The NETTER-2 trial compared ^177^Lu-Dotatate with high-dose long acting octreotide (60 mg monthly) as first-line systemic therapy in patients with higher-risk grade 2 NETs (Ki-67 ≥10%) and well-differentiated grade 3 NETs (Ki-67 >20% to ≤55%). Upfront PRRT yielded superior progression-free survival (the primary endpoint) to high-dose octreotide, 22.8 months vs. 8.5 months.^81^ Seventeen percent of the patients in the PRRT arm had peritoneal metastases but outcomes specific to that group were not reported specifically. Small bowel obstruction occurred in 5 of 147 patients (3.4%) receiving PRRT but none of the 73 patients on high-dose octreotide.

Recent data indicate that extensive peritoneal disease may be a partial contraindication to ^177^Lu-Dotatate. One retrospective study showed that PRRT did not control the peritoneal metastases in more than a third of the treated patients, and bowel obstruction was noted following PRRT.^5^ Another study described occurrences of bowel obstruction shortly after PRRT treatment, including two cases of frozen abdomen in patients with high-burden peritoneal carcinomatosis. The bowel obstruction may be caused by transient peritoneal inflammation following PRRT leading to peritoneal and/or mesenteric fibrosis.^23,63^ Corticosteroids have been proposed both for treatment as well as prophylaxis of radiation peritonitis, although their use has not been evaluated in prospective studies.

Additionally, the risk of bowel obstruction related to PRRT in patients with PM strongly correlates with both the number and size of peritoneal disease.^4,22^ Therefore, risks and benefits need to be carefully assessed prior to initiating PRRT, and the timing of surgery relative to PRRT should be addressed in a multidisciplinary setting. Patients with diffuse peritoneal carcinomatosis are likely poor candidates for treatment whereas patients with a small number of tumors are much more likely to tolerate treatment without obstructive complications.

### Chemotherapy

Cytotoxic chemotherapy is active in metastatic pancreatic NETs (30-40% response rate), however the role of chemotherapy in midgut NETs is more limited. Chemotherapy regimens such as capecitabine/temozolomide (CAPTEM) are associated with a very low response rate in grade 1 and 2 midgut NETs; however, one retrospective study indicated that response rate may be higher in grade 3 midgut NETs.^82^ FOLFOX and CAPEOX can also be considered in patients with high volume of disease or aggressive clinical progression.

### Systemic therapy for NECs

Cisplatin or carboplatin with etoposide remains the standard first-line regimen for metastatic poorly differentiated NEC but other regimens such as FOLFIRINOX may also be reasonable.^83^ The FOLFIRINEC trial (NCT04325425) is prospectively comparing cisplatin/etoposide with FOLFIRINOX as first-line therapy is ongoing. Given the highly aggressive nature of NEC, the role of surgery is very limited.

## DISCUSSION

Herein we summarize the updated consensus guidelines on the management of NENs with PMs. Our current consensus group expanded to include surgical oncologists, medical oncologists, radiologists, pathologists, and patient advocates. Consensus was achieved in all seven question blocks after two rounds of review. Despite the low level of evidence, substantial work had been produced in the field of NENs to require major adoptions and revisions.

The current approach improves upon the classification of NENs with PMs based on disease grading and symptomatology. Key recommendations from the consensus, which received strong support, include the early identification of carcinoid syndrome and its complications, such as carcinoid heart disease, as well as other functional tumors. Those at high risk of carcinoid heart disease should undergo cardiac monitoring. The administration of short-acting octreotide before CRS should be considered in patients with carcinoid syndrome in the setting of NET-PMs to avert carcinoid crisis, although the benefit of this routine practice has recently been called into question. Additionally, in G1 and G2 well-differentiated NETs with PMs, CRS is recommended if the burden of liver and distant metastases is low. This may include parenchymal-sparing hepatectomy or ablative techniques. Moreover, patients experiencing bowel obstruction secondary to PMs, often in the setting of carcinoid fibrosis, may require CRS or palliative surgical correction. Such cases necessitate multidisciplinary supportive care with a focus on symptom mitigation and social support. Lastly, systemic therapy is recommended as first-line treatment in scenarios involving progressive disease of grade 3 NETs or NECs. Recently, the CABINET trial showed that the use of cabozantinib in advanced extra-pancreatic and pancreatic NETs improved PFS compared to placebo.^84^

Major limitations of this expert consensus merit discussion. Firstly, the available evidence for our rapid review were of low quality and scarce. Therefore, the consensus methodology was employed to provide guidance regarding matters of equipoise. Secondly, the expert panel consisted primarily of surgical oncologists. Having expected this bias from the inception phases, thought leaders in medical oncology and other disciplines were involved early on for reviewing feedback from the first Delphi round and outlining principles of systemic therapy. Lastly, the Delphi consensus entailed voting on blocks rather than individual itemized recommendations, aligning with the original Chicago Consensus framework.^7,85^ While this approach helped mitigate survey fatigue, it may have compromised the granularity of feedback received.

### PSOGI: International Perspective

Management of NET-PMs is nuanced and understudied, as evidenced by guidelines from major multi-national organizations, such as European Society for Medical Oncology (ESMO), ENETs, NANETs, and APNETs.^33,86-89^ These conditions are rare, leading to limited specific recommendations. Current guidelines suggest surgery as a primary option for low-grade, low-burden NETs, yet less clarity is provided for NET-PMs. ESMO 2020 recommends surgery for local or locoregional G1 and G2 NETs, advocating for medical management or a watch-and-wait strategy in certain cases, such as those with a low Ki-67 index and stable disease. ENETs 2016 guidelines endorse surgery with curative intent if significant cytoreduction is achievable, aiming to avoid complications such as obstruction, fibrosis-induced events, external bowel compression, or portal hypertension.

Liver metastases, a frequent co-occurrence in NETs, are addressed in several guidelines. ENETs advises cytoreductive surgery for symptom control in functional NETs with liver-dominant disease, even if complete tumor removal is not possible. ESMO 2020 and Nordic 2014 guidelines recommend local treatments and systemic therapies in combination with surgery. Controversy remains regarding the extent of tumor removal necessary for debulking surgery, with recent shifts towards a debulking threshold of 70% from 90%.

In concordance with PSM guidelines, palliative resection of the primary tumor in advanced NETs is typically recommended to avoid complications such as bowel obstruction or ischemia, as per ESMO 2020 guidelines. However, the efficacy of primary tumor removal in improving survival in metastatic disease remains a topic of debate. NEC is highlighted as an absolute contraindication for upfront surgery, with such patients being directed towards systemic therapies or enrollment in clinical trials. Lastly, no clinical data currently exist for perioperative intraperitoneal chemotherapy or HIPEC in the setting of PMs in GEP-NETs.

### Patient and Caregiver Perspective

Dealing with NENs that have spread to the peritoneum presents significant challenges for patients and their loved ones. Concerns include the scarcity of clinical trials for advanced NENs and the exclusion of patients with PMs. This exclusion not only limits access to potentially beneficial treatments but also impedes progress in understanding this disease, leaving patients feeling marginalized and without options for advanced care. The difficulty in accurately assessing eligibility for trial enrollment using standard imaging techniques poses another barrier, culminating in uncertainty and anxiety among patients and caregivers about the best course of action.

Inclusion of patients with PM in clinical trials is especially important in addressing complications such as diffuse carcinomatosis, ascites, bile acid diarrhea, and malignant bowel obstruction. These conditions are particularly debilitating and can impact quality of life. Unfortunately, patients with malignant bowel obstruction are often doubly disadvantaged, being excluded from both oral antineoplastic therapies and intravenous (IV) treatments. Centers that are well-equipped with the necessary medical and psychosocial resources to facilitate a comprehensive, multidisciplinary approach are essential for effectively addressing the complexities of PM.

Patients have also emphasized the importance of using outcome measures addressing quality of life, palliation, and functional status alongside traditional metrics such as survival, complication rates, and response to therapy. Akin to the clinical trials guide provided by LACNETs, patients have expressed a need for informational handouts on managing a life with advanced NENs, including access to resources for mental and emotional health.

## CONCLUSION

In summary, we report an updated Delphi consensus on the management of NENs with peritoneal metastasis that included a multidisciplinary team of experts, including surgical/medical oncologists, radiologists, pathologists, and patient support groups. Cytoreductive surgery is highly recommended in cases of well-differentiated, low-grade NETs with treatable peritoneal and hepatic metastases. Higher grade, well differentiated disease typically undergoes systemic therapy considerations prior to consideration of cytoreduction. NECs are usually treated with up-front chemotherapy.

## Data Availability

The datasets used and/or analysed during the current study are available from the corresponding author on reasonable request.

