## Supplementary material for "Consensus Guideline for the Management of Peritoneal Metastases in Patients with Neuroendocrine Neoplasms": n/a

**Figure S1: Delphi Round 1 - Clinical Pathway for Management of Neuroendocrine Neoplasms with Peritoneal Metastases**

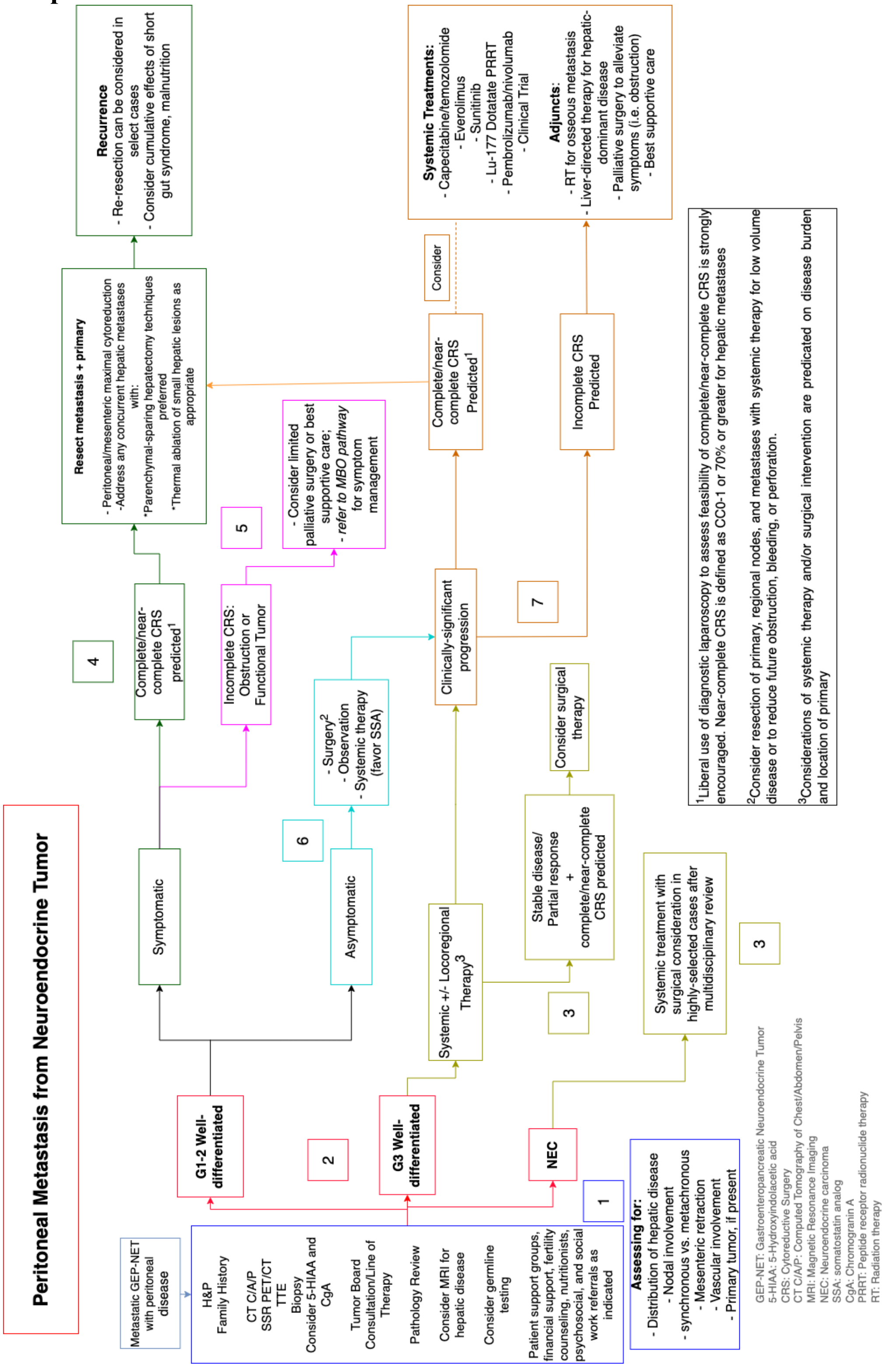

**Table S1: Quality Assessments**

| Author | Selection | Comparability | Outcomes | Overall |
|---|---|---|---|---|
| **Benhaim et al. [17]** | *** | - | *** | ****** |
| **Boer et al. [18]** | ** | - | *** | ***** |
| **Elias et al. [19]** | **** | - | ** | ****** |
| **Elias et al. [20]** | **** | - | *** | ******* |
| **Hajjar et al. [21]** | **** | - | *** | ******* |
| **Merola et al. [5]** | **** | - | *** | ******* |
| **Wonn et al. [4]** | *** | - | *** | ****** |
| **Wright et al. [22]** | *** | - | *** | ****** |
| **Strosberg et al. [23]** | *** | - | *** | ****** |

**Table S2: Search strategy for rapid review**

| | |
|---|---|
| 1 | periton*[TW] OR carcinomatosis[tw] OR "peritoneal neoplasms"[Mesh Major topic] OR HIPEC OR intraperitoneal chemotherapy[TW] OR "Hyperthermic intraperitoneal chemotherapy"[Mesh Major Topic] OR (periton*[tw] AND cytoreduc*[TW]) |
| 2 | "neuroendocrine tumor"[tw] OR "neuroendocrine tumors"[Mesh Major Topic] OR carcinoid*[tw] OR "Receptors, Peptide"[MeSH Major Topic] OR "peptide receptor"[tw] OR PRRT[tw] OR DOTATE[tw] |
| 3 | #1 and #2 |
